# Higher frequency of interstate over international transmission chains of SARS-CoV-2 virus at the Rio Grande do Sul - Brazil state borders

**DOI:** 10.1101/2024.05.21.24307668

**Authors:** Filipe Zimmer Dezordi, José Valter Joaquim Silva Júnior, Terimar Facin Ruoso, Angela Giovana Batista, Pedro Mesquita Fonseca, Richard Steiner Salvato, Tatiana Schäffer Gregianini, Thaísa Regina Rocha Lopes, Eduardo Furtado Flores, Rudi Weiblen, Patrícia Chaves Brites, Mônica de Medeiros Silva, João Batista Teixeira da Rocha, Gustavo de Lima Barbosa, Lais Ceschini Machado, Alexandre Freitas da Silva, Marcelo Henrique Santos Paiva, Matheus Filgueira Bezerra, Tulio de Lima Campos, Tiago Gräf, Daniel Angelo Sganzerla Graichen, Elgion Lucio da Silva Loreto, Gabriel da Luz Wallau, Fiocruz Genomic Network

**Affiliations:** Departamento de Entomologia, Instituto Aggeu Magalhães (IAM)-Fundação Oswaldo Cruz-FIOCRUZ, Recife 50670-420, Brazil; Núcleo de Bioinformática (NBI), Instituto Aggeu Magalhães (IAM), FIOCRUZ-Pernambuco, Recife, Pernambuco, Brazil; Setor de Virologia, Departamento de Medicina Veterinária Preventiva, Universidade Federal de Santa Maria (UFSM), 97105-900, Santa Maria, RS, Brazil; Setor de Virologia, Instituto Keizo Asami, Universidade Federal de Pernambuco,Pernambuco, Brazil. Zip Code: 50670-901; Laboratório NB3 de Neuroimunologia, Universidade Federal de Santa Maria, Rio Grande do Sul, Brazil. Zip Code: 97105-900; Departamento de Microbiologia e Parasitologia, Universidade Federal de Santa Maria, Rio Grande do Sul, Brazil. Zip Code: 97105-900; Programa de Pós-graduação em Medicina Veterinária, Universidade Federal de Santa Maria, Rio Grande do Sul, Brazil. Zip Code: 97105-900; Campus Palmeira das Missões, Universidade Federal de Santa Maria. Palmeira das Missões, Rio Grande do Sul, Brazil. Zip Code: 98300-000; Life Sciences Institute, Universidade Federal de Juiz de Fora. Governador Valadares, Minas Gerais, Brazil. Zip Code: 35010-180; Centro Estadual de Vigilância em Saúde. Secretaria Estadual da Saúde do Rio Grande do Sul. Porto Alegre, Rio Grande do Sul, Brazil. Zip Code: 90610-000; Hospital Universitário de Santa Maria (HUSM), Universidade Federal de Santa Maria (UFSM), Av. Roraima, 1000, 97105-900, Santa Maria, RS, Brazil; Departamento de Bioquímica e Biologia Molecular, Universidade Federal de Santa Maria (UFSM), Av. Roraima, 1000, 97105-900, Santa Maria, RS, Brazil; Núcleo de Plataformas Tecnológicas (NPT), Instituto Aggeu Magalhães (IAM), FIOCRUZ-Pernambuco, Recife, Pernambuco, Brazil; Núcleo de Ciências da Vida, Universidade Federal de Pernambuco (UFPE), Centro Acadêmico do Agreste-Rodovia BR-104, km 59-Nova Caruaru, Caruaru 55002-970, Brazil; Departamento de Microbiologia, Instituto Aggeu Magalhães (IAM), FIOCRUZ-Pernambuco, Recife, Pernambuco, Brazil; Laboratório de Virologia Molecular, Instituto Carlos Chagas, Fundação Oswaldo Cruz, Curitiba, Paraná, Brazil; Departamento de Zootecnia e Ciências Biológicas, Universidade Federal de Santa Maria, Palmera das Missões, Rio Grande do Sul, Brazil. Zip Code: 98300-000; Department of Arbovirology, Bernhard Nocht Institute for Tropical Medicine, WHO Collaborating Center for Arbovirus and Hemorrhagic Fever Reference and Research. National Reference Center for Tropical Infectious Diseases. Bernhard-Nocht-Straße 74, 20359 Hamburg - Germany

## Abstract

Brazil’s COVID-19 response has faced challenges due to the continuous emergence of variants of concern (VOCs), emphasizing the need for ongoing genomic surveillance and retrospective analyses of past epidemic waves. Rio Grande do Sul (RS), Brazil’s southernmost state, has crucial international borders and trades with Argentina and Uruguay, along with significant domestic connections. The source and sink of transmission with both national and international hubs raises questions about the RS role in the transmission of the virus, which has not been fully explored. Nasopharyngeal samples from various municipalities in RS were collected between June 2020 and July 2022. SARS-CoV-2 whole genome amplification and sequencing were performed using high-throughput Illumina sequencing. Bioinformatics analysis encompassed the development of scripts and tools to take into account epidemiological information to reduce sequencing disparities bias among the regions/countries, genome assembly, and large-scale phylogenetic reconstruction. Here, we sequenced 1,480 SARS-CoV-2 genomes from RS, covering all major regions. Sequences predominantly represented Gamma (April-June 2021) and Omicron (January-July 2022) variants. Phylogenetic analysis revealed a regional pattern for transmission dynamics, particularly with Southeast Brazil for Gamma, and a range of inter-regional connections for Delta and Omicron within the country. On the other hand, international and cross-border transmission with Argentina and Uruguay was rather limited. We evaluated the three VOCs circulation over two years in RS using a new subsampling strategy based on the number of cases in each state during the circulation of each VOC. In summary, the retrospective analysis of genomic surveillance data demonstrated that virus transmission was less intense between country borders than within the country. These findings suggest that while non-pharmacological interventions were effective to mitigate transmission across international land borders in RS, they were unsuficient to contain transmission at the domestic level.

## 1. Introduction

The first cases of effective human-to-human transmission of SARS-CoV-2 were identified in Wuhan, China, in December 2019 (1). Since then, this virus has spread globally culminating in a pandemic declared on 11th March 2020 (2). As of February 2024, there have been 676 million global cases and more than 6.8 million deaths (3) and Brazil registered more than 37 million of the total cases and almost 700 thousand deaths in the same period (3). These figures are a consequence of the high transmissibility of air-born pathogens such as the SARS-CoV-2 through symptomatic and asymptomatic patients and continuous adaptations to the new human host (4–6). The rapid evolution of the SARS-CoV-2 under changing evolutionary pressures associated with millions of infected patients (3) has resulted in the emergence and spread of more transmissible and/or more immunological evasive SARS-CoV-2 lineages around the globe that are dynamically classified as Variants of Interest (VOIs) and Variants of Concern (VOCs) (7).

Besides the development of different vaccine technologies (8) and the success of vaccine campaigns across the world, which culminated in a decrease in cases, particularly in severe disease and hospitalizations (9–11), the SARS-CoV-2 is still circulating and evolving into new lineages. These new lineages carry specific sets of immune escape mutations allowing the reinfection of previously infected or vaccinated hosts (12–14). Since the start of the pandemic, five major VOCs encompassing several lineages have originated and spread across the globe. The first VOC identified was the Alpha lineage in September 2020 (15) followed by Beta in October 2020 (16), Gamma in November 2020 (17), Delta in October but spread globally in May 2021 (18), and Omicron in November 2021 (19), which is the single VOC circulating nowadays, and now considered as a VOI (20), with several sublineages co-circulating (20).

Knowledge about the spreading of different SARS-CoV-2 lineages is only possible due to the efforts of genomic surveillance, an effort carried out by thousands of research groups across the globe (21,22). In Brazil, due to its large geographic territory, several groups have been characterizing the emergence and spreading of different SARS-CoV-2 lineages across the country (17,23–27). Besides the efforts to perform genomic surveillance studies in Brazil, the majority of Brazilian studies focus on the Southeast region - the most populous region, and with the major number of cases - (25). Moreover, the studies focused on Rio Grande do Sul were carried out at the beginning of the pandemic (28–30) or focused on specific cities (31,32). Herein, we aimed to characterize the circulation of SARS-CoV-2 lineages during the first two and a half years of the pandemic (up to July 2022) across Rio Grande do Sul state state which have epidemiological links with samples from other Brazilian states, besides the large land border of Rio Grande do Sul with Argentina and Uruguay.

We sequenced 1,480 genomes from samples collected from June 2020 to July 2022. The genomes represent several lineages from Gamma, Delta, and Omicron VOCs as well as non-VOI and non-VOC lineages. The samples represent different cities across the Rio Grande do Sul. These findings are of particular importance to prioritize non-pharmacological interventions at the interstate and national level during the following SARS-CoV-2 waves.

## 2. Material and Methods

Nasal and oropharyngeal specimens, previously identified as positive for SARS-CoV-2, were obtained from the Universidade Federal de Santa Maria main campus and the Palmeira das Missões Campus representing samples from 277 Rio Grande do Sul municipalities mainly covering the Central and Northwest regions of the state during the period from 23 June 2020 to 7 July 2022. Reverse Transcription Quantitative Polymerase Chain Reaction (RT-qPCR) assays were conducted utilizing the Biomol OneStep Covid-19 Kit (IBMP, Paraná, BR) and the Molecular SARS-CoV-2 Kit (Bio-manguinhos, Rio de Janeiro, RJ, BR), strictly adhering to the specifications provided by the manufacturers. Specimens exhibiting a Cycle Threshold (Ct) value below 25 were subsequently subjected to amplification and sequencing processes, as delineated in (33,34).

### 2.1. Viral Genetic Material Sequencing

For this study, two distinct methodologies were employed to synthesize complementary DNA (cDNA) and to amplify the SARS-CoV-2 genome. These included: the short amplicon approach based on the ARTIC protocol (accessible at https://github.com/artic-network/artic-ncov2019) (33); and the incorporation of three sets of primers within the COVIDSeq protocol, following Naveca, et al., 2021 (35). After the generation of genome-wide amplicons, samples were processed for sequencing employing either the Illumina DNA Prep (Illumina, San Diego, CA, USA) or the COVIDSeq (Illumina, San Diego, CA, USA) library preparation protocols, following the guidelines provided by the manufacturer. The sequencing process was executed using the Illumina MiSeq system, specifically employing the MiSeq Reagent Kit V3 for a paired-end 150 cycles flow cell. Ethical clearance for this study was granted by CEP UFSM under the authorization number 52939821.5.0000.5346 and 47588621.7.1001.5346 and by CEP from Escola de Saúde Pública/SES-RS under the authorization number CAAE: 67181123.1.0000.5312.

### 2.2. Genome Assembly

In this study, we used the ViralFlow v0.0.6 workflow (36). This workflow was designed for efficient and accurate assembly of viral genomes. Briefly, raw sequence data were quality filtered to remove reads (*--cut_front --cut_tail --qualified_quality_phred 20 -l 35 -f 35 -t 35 -F 35 -T 35*) with fastp tool (37). Subsequently, the cleaned reads were mapped with BWA (38) against the Wuhan SARS-CoV-2 genome (NC_045512.2). The consensus genomes were generated with the iVar tool (39), where a base is called to the consensus when it reaches at least 5x of coverage depth; on multi-allele loci, alleles with higher frequency (VAF >= 0.51) are considered for the consensus genome. The assembly metrics and SARS-CoV-2 lineages are defined using bamdst (40) and pangolin (41), respectively, both present in the ViralFlow workflow. All samples with 70% or more of coverage breadth are included in the initial phylogenetic analysis with MAPLE v0.3.1 (42) to detect if the lack of sequencing regions results in long branches on phylogenetic trees.

### 2.3. Subsampling Strategy

In light of the extensive dataset of SARS-CoV-2 genomes available in the GISAID-EpiCoV database (43), which encompasses millions of genomes globally (15,271,031 genomes in the collection date of this study), including hundreds of thousands from Brazil (214,213 genomes), it became imperative to utilize a systematic subsampling strategy informed by epidemiological data. To facilitate this, we developed an in-house R (https://www.r-project.org/) script, named explore.R (see **Data Availability** for details), to determine the appropriate number of genomes to subsample across various Brazilian regions for Gamma, Delta, and Omicron. This script harnesses data from both the EpiCoV database and covid19.org.br, collected on February 6, 2023, to assess and use the ratio of COVID-19 cases to estimate the proportion subsampling of the available genomes for each aforementioned VOCs in specific periods.

To delineate the temporal and spatial distribution of the Gamma, Delta, and Omicron variants within Brazil, we referred to existing literature to establish the onset of their circulation in the country: Gamma in November 2020 (17), Delta in April 2021 (18), and Omicron in November 2021 (19). Our study’s scope was extended until August 2022, based on the date of the most recent sample sequenced in our analysis.

To perform the subsampling process, we developed an in-house tool: the Gisaid Subsampling Toolkit, or GIST. GIST processes a JSON file containing the count of genomes from various Brazilian regions, derived from the output of the ‘explore.R’ script. It employs a combination of Augur (44), BLAST (45), and MAFFT (46) for three distinct analyses (**Code Availability** section for more details, **SupplementaryData1**). Briefly, in the first analysis, we utilize Augur to sample genomes. This step involves the exclusion of genomes from non-human hosts, those lacking a collection date, and sequences shorter than 28,400 bases, approximately 95% of the SARS-CoV-2 Wuhan genome’s coverage. The sampling is stratified based on the number of genomes by state, further grouped by pango lineage, year, and month. Following this initial sampling, the second phase of analysis enriches the dataset with genetically similar genomes. This is achieved through a BLAST analysis filtering results between 99% and 99.98% of identity with query coverage high scoring pais (HSP) greater or equal to 99.9%. In the final stage of analysis, we conduct a multiple sequence alignment using MAFFT, employing parameters such as --keeplength, --kimura 1, and --6merpair, while specifically masking the UTR regions.

### 2.4. Phylogenetic Analysis

Three initial trees - one for Gamma, Delta, and Omicron datasets - were reconstructed using MAPLE v0.3.1 (42) to detect long branches. Genomes showing long branches (**SupplementaryData2**) were removed from the original alignment. The edited alignments were used in a maximum likelihood phylogenetic analysis with IQ-TREE2 v.2.2.0 (47), the best substitution model was defined using ModelFinder (48) implemented on IQ-TREE2 and the SH-aLRT test (49) to branch support. The phylogenetic trees were annotated in iTOL v.6 (50) to identify clades of specific lineages with branch support higher than 0.8 (aLRT) with at least 4 genomes from Rio Grande do Sul. The identified clades were split into different datasets, one for each clade lineage.

### 2.5. Temporal Estimation

To estimate the tMRCA of clades identified in phylogenetic analysis each lineage dataset was used in an analysis flow which encompasses: A first phylogenetic tree reconstruction with IQ-TREE2 v.2.2.0; A TempEst v.1.5.3 (51) analysis to identify the age root correlation; removal of outliers with in-house R scripts, that receives the output of TempEst and identify outliers sequences based on the thresholds of 1.5x of Interquartile Range IQR of all samples in the dataset; A second tree reconstruction with IQ-TREE2 v.2.2.0 using the dataset alignments without outliers; And finally a temporal analysis with TreeTime v.0.11 (52).

## 3. Results

### 3.1. Sequencing Results

During the period and for the purpose of the present study, 1,480 genomes were sequenced and deposited on EpiCoV - GISAID database (**SupplementaryData3**). The samples contain representatives of most parts of the North, Center, and West of the Rio Grande do Sul state (https://microreact.org/project/2utyqQKoA7zDkYHEuzvyTG-sarsufsmlacen), and encompass approximately 30% of all genomes submitted for the state (1,480 from 5,065 genomes) and deposited on GISAID in the period of the study (**SupplementaryData3**). The submitted consensus genomes from our group represent samples that meet one of the following criteria: 90% or more of genome coverage breadth (horizontal coverage) or 70% or more genome coverage breadth and that do not belong to clades with long branches on initial MAPLE phylogenetic analysis. The samples are majorly composed of Gamma lineages and sublineages during the period from April to June 2021 (**Figure 1**), and Omicron lineages and sublineages during the period of January to July 2022, which is in line with the general numbers presented on EpiCoV - GISAID (**Figure 2**). Besides the analysis of this study is focused on SARS-CoV-2 samples sequenced in Rio Grande do Sul, we noticed similar patterns of lineage frequencies sequenced in other regions of Brazil (**Figure 2**). The most prevalent lineages for each analyzed VOC on Rio Grande do Sul were P.1, P.1.14, P.1.2, P.1.4 and P1.7 for Gamma; AY.43, AY.99.2, AY.99.1, AY.101, AY.34.1 and AY.46.3 for Delta; and the most prevalent lineages BA.1, BA.1.1, BA.2, BA.5, BA.5.2, BA.4 and BQ for Omicron (**Figure 2**).

**Figure 1:**
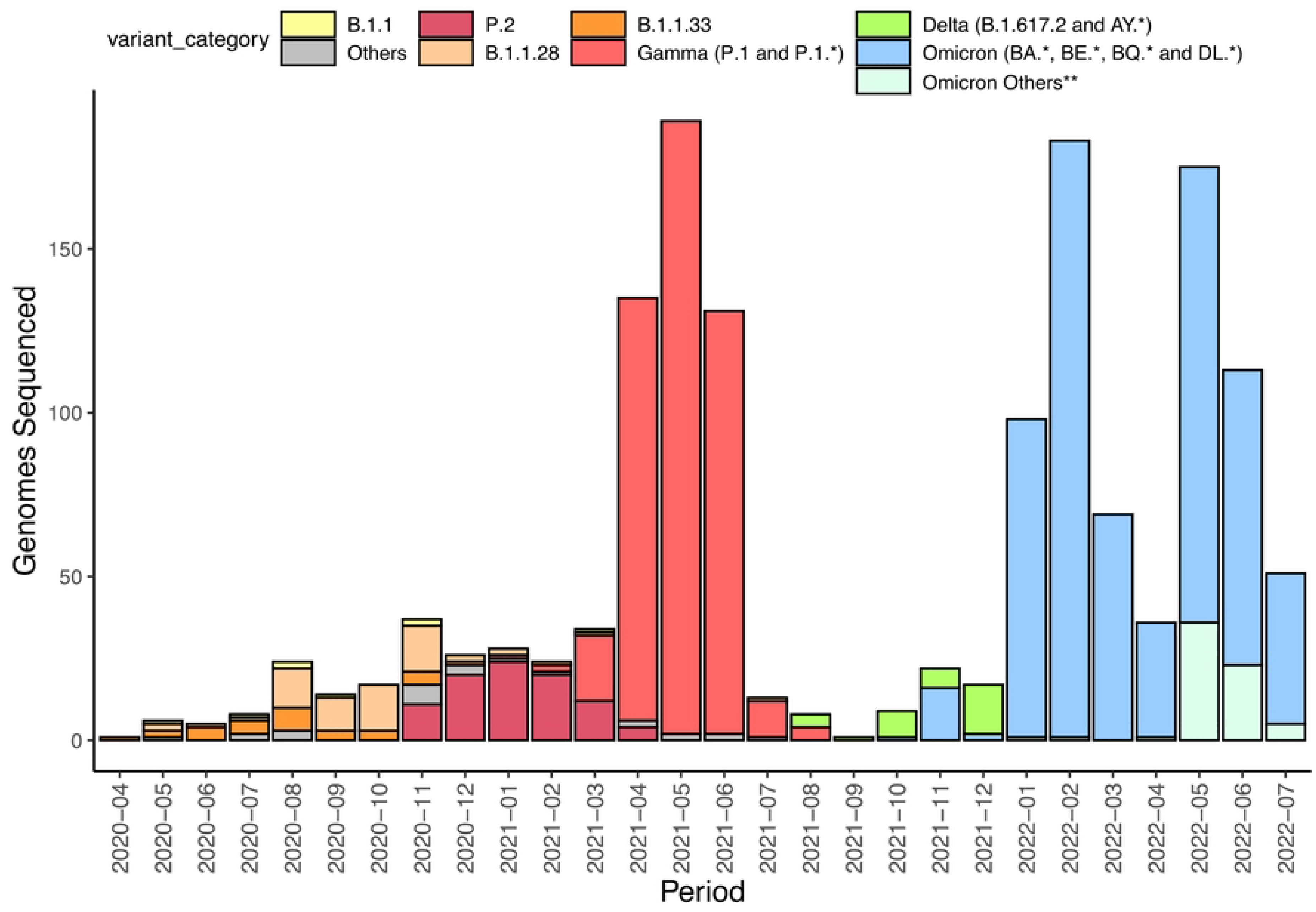
Number of samples sequenced in this study per month. **Omicron genomes not related to BA, BE, BQ, and DL lineages.

**Figure 2.**
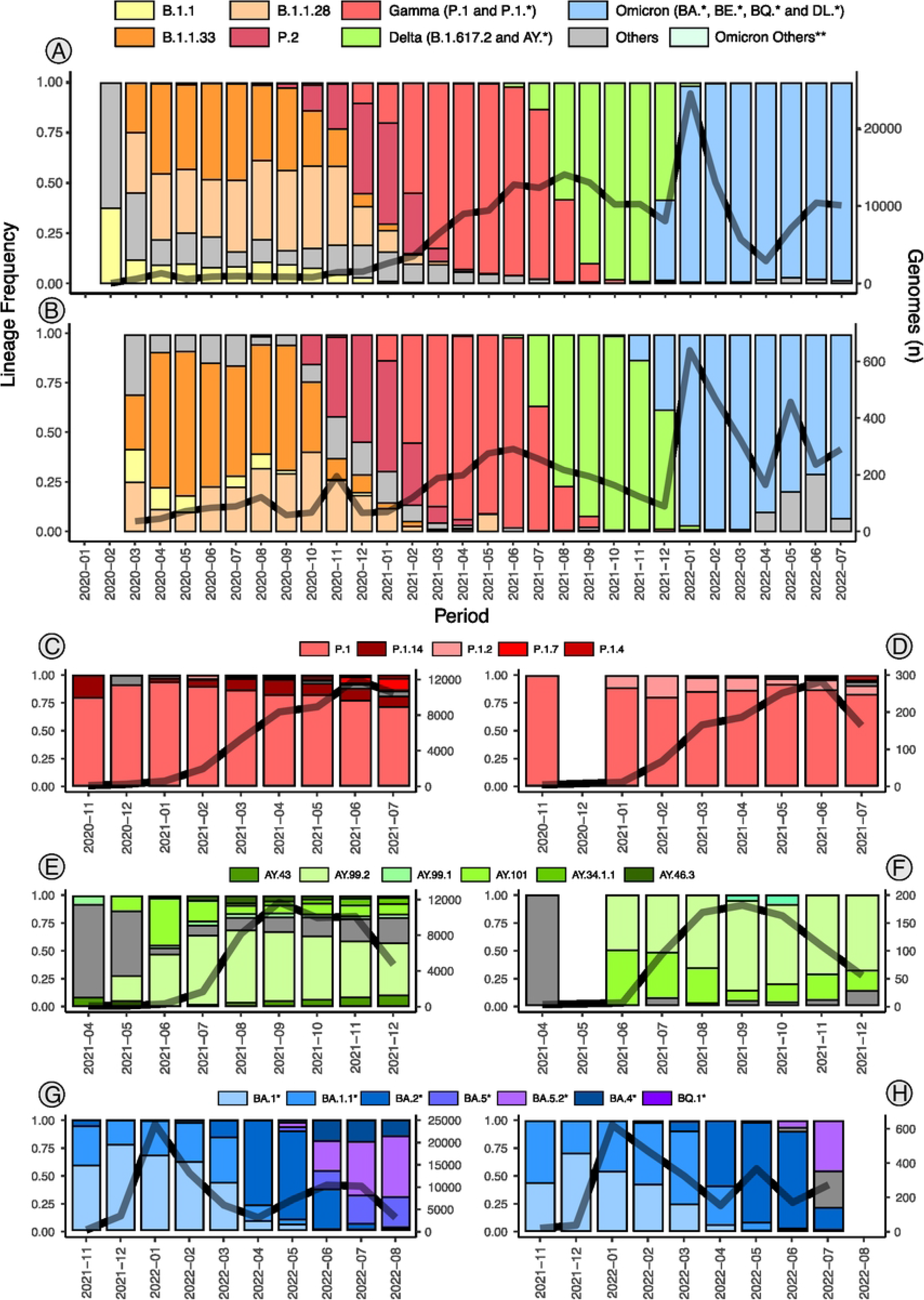
Prevalence of SARS-CoV-2 lineages and number of genomes per period, based on EpiCoV GISAID data. **A**. Specific lineages and variants in Brazil. **B**. Specific lineages and variants in Rio Grande do Sul. **C**. Gama lineages in Brazil. **D**. Gama lineages in Rio Grande do Sul. **E**. Delta lineages in Brazil. **F**. Delta lineages in Rio Grande do Sul. **G**. Omicron lineages in Brazil. **H**. Omicron lineages in Rio Grande do Sul. **Omicron genomes not related to BA, BE, BQ, and DL lineage.

### 3.2. SARS-Cov-2 clades circulating on Rio Grande do Sul

The subsampling strategy with explorer.R and GIST resulted in three major datasets: One for each VOC Gamma, Delta, and Omicron, encompassing the most frequent lineages on Rio Grande do Sul during the period of the study as well as outgroups with ancestors of respective lineages (**SupplementaryData4**). Each dataset corresponds to Brazilian genomes plus genomes from Latin America (particularly Paraguay, Argentina and Uruguay) and other regions of the globe, recovered from the GIST BLAST analysis (**SupplementaryData5**).

The initial phylogeny of Gamma corresponds to 4,545 genomes (**Figure 3A**), where 6 clades were identified (**Figure 3B**). One clade (P.1.14-I) represents sequences from P.1.14, two clades (P.1-I∼II) represent sequences of P.1 lineage, and three (P.1.2-I∼III) of P.1.2 lineage. The Gamma analysis shows samples from Rio Grande do Sul clustering with sequences from the Southeast region of Brazil: The P.1.14-I and P.1-II clades encompass basal sequences from Minas Gerais (MG) and São Paulo (SP) states, the P.1-I clade presents basal Rio Grande do Sul samples clustered with South America samples from Argentina and Paraguay, and the clades P.1-II and P.1.2-III have well-defined sister clades from Uruguay and São Paulo, respectively.

**Figure 3.**
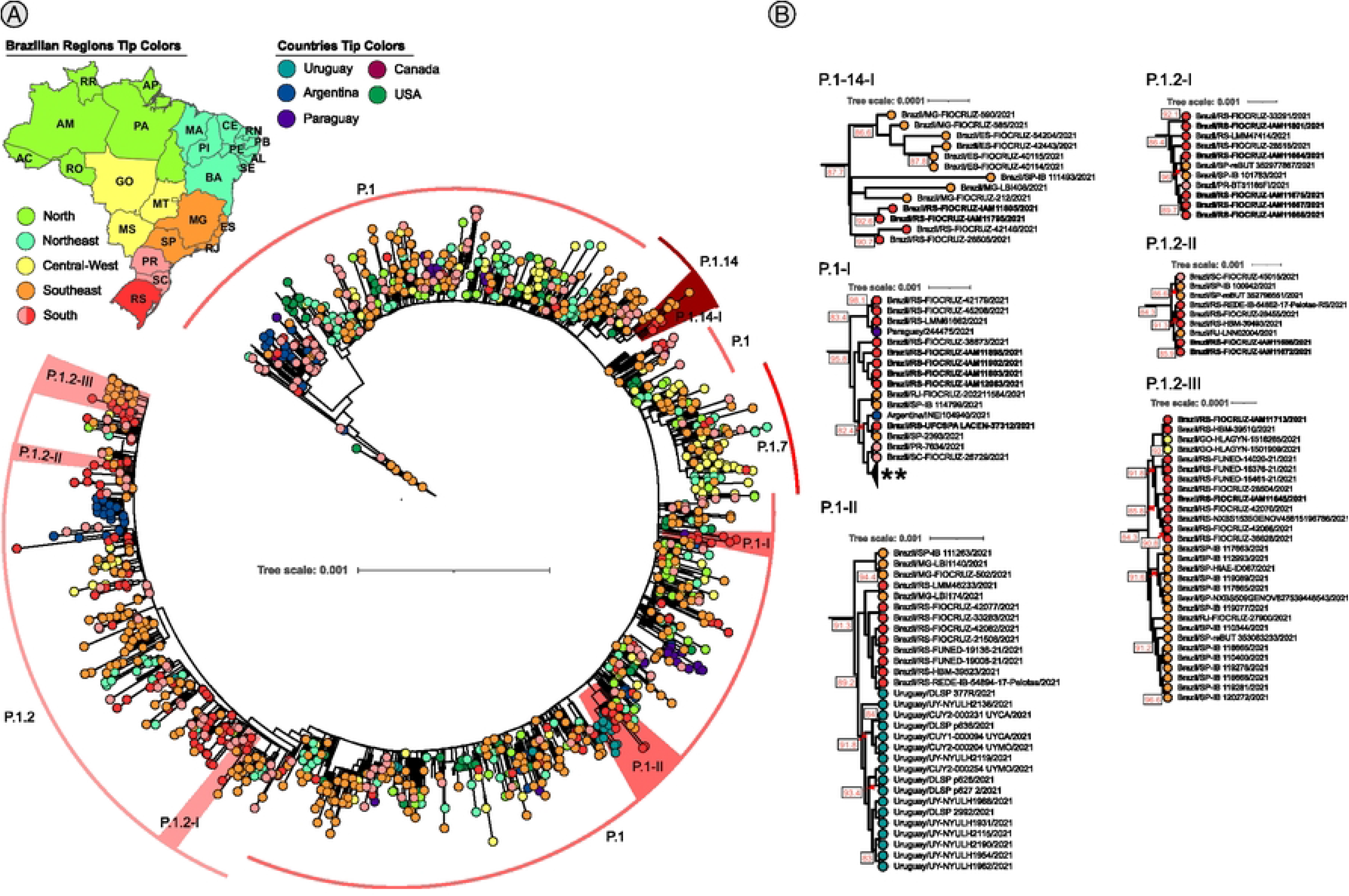
Maximum likelihood analysis of Gamma variant lineages, Branch support was accessed by the aLRT branch support. Tip colors are based on Brazilian regions and the 5 countries that presented the highest number of sequences recovered via blast. **A**. Complete tree. **B**. Well-supported subclades. Bold sample lanes represent genomes sequenced in this study. ** Collapsed clade that represents all sequences after P.1-I clade. Complete figure with branch support and uncollapsed clade is available on **SupplementaryData6**.

The initial phylogeny of Delta corresponds to 4,358 genomes (**Figure 4A**) where 6 clades were identified (**Figure 4B**). Four clades (AY.99.2-I∼IV) represent sequences from AY.99.2 lineage, and 2 clades (AY.101-I∼II) represent sequences from AY.101 lineage. The Delta analysis shows different clustering of Rio Grande do Sul (RS) and sampled from other Brazilian regions. The AY.99.2-I clade shows a possible transmission from Rio Grande do Sul to Minas Gerais, and the AY.99-2-IV from Minas Gerais to Rio Grande do Sul. The clade AY.101-I shows a route of transmission from Rio Grande do Sul to the Central-West region of Brazil, as well as from Rio Grande do Sul to Rio de Janeiro (RJ), and the AY.101-II clade from Rio Grande do Sul to São Paulo state.

**Figure 4.**
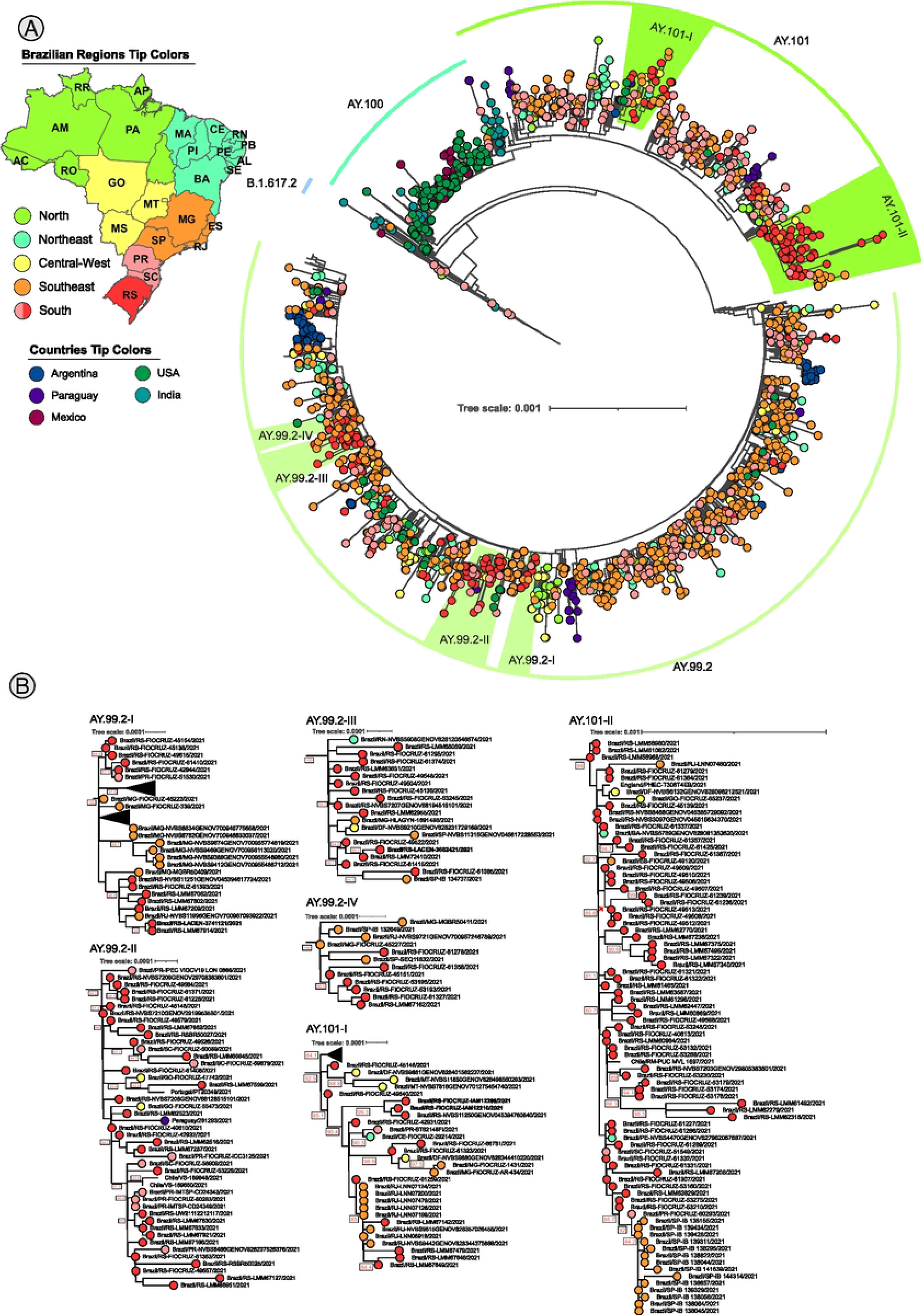
Maximum likelihood analysis of Delta variant lineages, numbers within with squares are aLRT branch support of respective clades. Colorful tips based on Brazilian regions and the 5 countries that presented the highest number of sequences recovered via blast. **A**. Complete tree. **B**. Weel supported subclades including Rio Grande do Sul samples. Bold sample lanes represent genomes sequenced in this study. Complete figure with branch support and uncollapsed clade is available on **SupplementaryData7**.

The omicron phylogenetic analysis was performed with 4,639 genomes (**Figure 5A**) representing 5 distinct clades (**Figure 5B**) with Rio Grande do Sul samples from BA.1.1, BA.5.2, and BA.2 lineages. Different from Delta and Gamma analysis, where several clades clustered with other clades from the South-East region - mainly from São Paulo, Rio de Janeiro, and Minas Gerais states - the Omicron clades are mostly composed of Rio Grande do Sul samples, except for BA.5.2-I clade, that clustered with samples from the Goias state (GO) - Central-West region.

**Figure 5.**
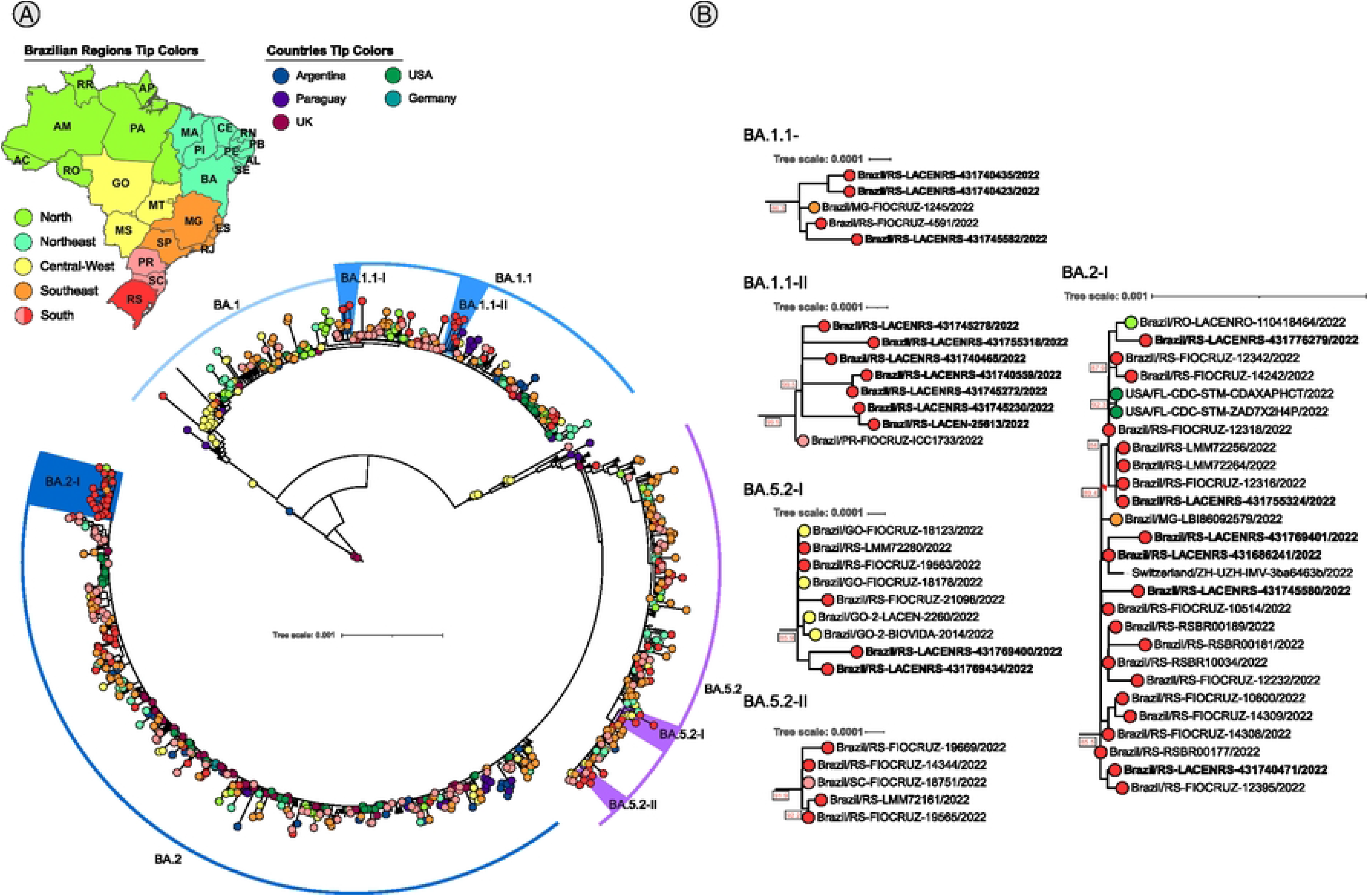
Maximum likelihood analysis of lineages of the Omicron variant, aLRT branch support. Colorful tips based on Brazilian regions and the 5 countries that presented the highest number of sequences recovered via blast. **A**. Complete tree. **B**. Subclades. Bold sample lanes represent genomes sequenced in this study. Complete figure with branch support and uncolapsed clade is available on **SupplementaryData8**.

### 3.3. Temporal estimation of 7 lineages circulating on Rio Grande do Sul

To identify the possible emergence and spreading timing of the lineages associated with the identified clades within the state, we performed temporal analysis using treetime for 7 datasets (**Table 1**) which represents the clades identified with Rio Grande do Sul samples in the evolutionary analysis.

**Table 1.** Datasets and tMRCA inferred by treetime tool.

| lineage | number of samples | tMRCA |
| --- | --- | --- |
| P.1.2 | 482 | 2021-01-12 ~ 2021.03 |
| P.1.14 | 540 | 2020-12-15 ~ 2020.95 |
| AY.101 | 1030 | 2021-03-22 ~ 2021.22 |
| AY.99.2 | 1953 | 2021-03-26 ~ 2021.23 |
| BA.1.1 | 1467 | 2021-10-09 ~ 2021.77 |
| BA.5.2 | 384 | 2022-02-11 ~ 2022.11 |
| BA.2 | 1530 | 2021-12-09 ~ 2021.93 |

Considering the tRMCA from treetime as the start time of lineages circulation, and the last collection date of samples (**Supplementary Data3**) as the end time of circulation, the Gamma P.1.2 and P.1.14 lineages were estimated to have occurred on Rio Grande do Sul between mid-December 2020 to late June 2021 for P.1.14 and mid-January 2020 to late June 2021 for P.1.2.

For Delta lineages, the AY.101 was estimated to have circulated between mid-March 2021 to late November 2021 and the AY.99.2 from late March 2021 to early December 2021, and for Omicron lineages, the BA.1.1 started circulating in the state in mid-August 2021, the BA.2 in mid-December 2021 and the BA.5.2 in mid-February 2022 and both of them are still circulating during the time of the data collection of this study: early July 2022.

## 4. Discussion

The SARS-CoV-2 virus is the latest example of a zoonotic pathogen that has made its way to effective human-to-human transmission resulting in the COVID-19 pandemic. Despite numerous efforts to implement large-scale genomic surveillance around the world, there remain underrepresented populations and areas where laboratory and genomic surveillance is scarce (21). In Brazil, SARS-CoV-2 genomic surveillance was established across the country, but as expected, most of the data comes from the most populated cities on the Atlantic coast and large capital cities from states without sea borders where there were more available diagnostic and research laboratories. That is the case of the Rio Grande do Sul state where there is a higher concentration of diagnostic and genomic information from the capital city Porto Alegre and surrounding municipalities. In this study, we generated 1,480 genomes mainly from the central and Northeast regions of the state to augment the genomic information from the Rio Grande do Sul state. Then we subsampled the main lineages circulating in the state to test hypotheses regarding the sink and source role of the state to other Brazilian states and border countries Argentina and Uruguay. We found that inter-state transmission was much more important in SARS-CoV-2 lineages transmission spread from and to the Rio Grande do Sul state than inter-country transmission.

The extremely large number of SARS-CoV-2 genomes currently available (the global dataset used in this study included 15,271,032 genomes) makes phylogenetic inferences timing and computationally consuming (53). Current approaches to handle such a massive amount of data are focused on lineages subsampling taking into consideration lineages presence in different locations and prevalence (54). The Augur software (44), implements some sub-sampling algorithms but does not consider the epidemiological estimates such as the number of cases per region. Considering the global disparities of genome sequencing throughout different geographic regions (21) we implemented a sampling strategy based on the number of cases and genomes by region per time using the in-house script explore.R which allowed us to more precisely sample genomes following the increase and decrease in cases and remove the sampling biases present in the global and local datasets.

Our global subsampled phylogenetic reconstruction of Gamma and Delta clades showed that the United States of America and the Central-West region of Brazil were placed in a basal position near the root suggesting that these regions were the source point to spread these lineages to Brazil. Previous evidence has shown the origins of each analyzed VOC, such as Gamma with P.1 lineage in the North region of Brazil (17), Delta with B.1.617.2 lineage in India (18), and Omicron with B.1.1.529 lineage in South Africa (19). But this is the first large-scale analysis showing that the Delta lineage was probably introduced from the USA to Brazil and that the Central-West region of Brazil acted as an important hub for the spreading of the Omicron lineage to other major regions of the country.

Based on the augmented dataset of SARS-CoV-2 genomes from Rio Grande do Sul including genomes of previously undersampled regions (Center and Northwestern) we next evaluated if the main transmission chains with Rio Grande do Sul occurred with other Brazilian states or with border countries Argentina and Uruguay. Previous studies characterized the spreading of B.1.1.28 and B.1.1.33 lineages from Rio Grande do Sul to Uruguay (55). Besides the large terrestrial frontier between Rio Grande do Sul and Uruguay and Argentina, our results showed only one probable migration event of the P.1 lineage from Rio Grande do Sul to Uruguay (**Figure 3B**, clade P.1-II). The clades of P.1.2 lineage showed samples from Rio Grande do Sul as ancestors from samples of Southeast, in line with the evidence showing the likely origin of this sub-lineage in the state (31). The P.1.14-I clade, showed an ancestral origin of P.1.14 samples from Southeast towards Rio Grande do Sul in line with the findings shown by Ferrareze, et al. 2024 (56).

There are few studies about Delta lineages on Rio Grande do Sul, one of the reasons may be related to the low number of samples sequenced in this period (**Figure 2B**). Our results showed that two main Delta lineages circulated on Rio Grande do Sul: the AY.99.2 and the AY.101, corroborating the results from Castro et al, 2023 (57) and Arantes et al, 2022 (27) that addressed the early emergence of Delta in Brazil. The circulation of those lineages appears to have occurred from July 2021 to January 2022, with the tMRCA of those lineages estimated from late March 2021, which is in line with the estimations of Arante et al, 2023 for AY.101 (27). The clades of AY.99.2 showed samples from Rio Grande do Sul as ancestors (**Figure 4B** Clade AY.99.2-I) with spreading to Minas Gerais (Southeast region), while the clade AY.99.2-IV showed the opposite, which indicates a spreading of AY.99.2 between those 2 states. The analysis of AY.101 clades also revealed Rio Grande do Sul samples as ancestors and a putative migration to Southeast states: Rio de Janeiro (AY.101-I clade) and São Paulo (AY.101-II clade).

Our results showed that ancestral samples of AY.99.2 from the Rio Grande do Sul state and posterior spreading to the Southeast were in line with other studies (56). On the other hand, the same study identified the introduction of AY.101 lineage in the Southeast region in early January 2021, and the posterior introduction in Rio Grande do Sul in July 2021 which diverged from our findings of an introduction from Rio Grande do Sul to the Southeast region. The divergence about the AY.101 phylogeographic inferences may be the result of different subsampling strategies, where Ferraze et al, 2024 (56) used the Augur subsampling strategy based on Pango lineage, and we used the Augur inside GIST performing a subsampling based on normalized genomes by state cases and then the sampling by Pango Lineage and collection date.

The Omicron analysis showed three main lineages circulating on Rio Grande do Sul during the period of the study: BA.1.1, BA.5.2, and BA.2 which is in line with other Brazilian states (**Figure 2G, H**). During the first quarter of 2022, the BA.1 and BA.1.1 lineages were the most prevalent, as described previously (58) but were further replaced by BA.2, BA.5, and BQ from June to July 2022 (59). There is no phylogenetic information in the literature about the relation of Omicron lineages from Rio Grande do Sul and other locations, our results showed that excluding the BA.5.2-I clade (**Figure 5B**), the omicron clades are majorly formed by samples from Rio Grande do Sul. The BA.5.2-I clade showed a close relation between Rio Grande do Sul samples and Goias samples (Central West Brazilian region).

Despite the standardization of sampling methods, sequencing disparities (21) may still generate sampling biases that can affect genomic surveillance studies, here we identified both local sampling disparities on the Rio Grande do Sul (by state regions and periods, see https://microreact.org/project/2utyqQKoA7zDkYHEuzvyTG-sarsufsmlacen) and in countries that have terrestrial borders with the Southern region of Brazil (Paraguay, Argentina, and Uruguay, see **Supplementary Data 9**). These results may have biased our inferences, but on the other hand, this was the most comprehensive study posed to augment the Rio Grande do Sul dataset in underrepresented areas as well as to employ a case incidence aware subsampling scheme that minimized as much as possible the impact of these biases as much as possible.

Overall, our findings suggest that despite the large land border and intense international transportation of goods between Rio Grande do Sul, Argentina, and Uruguay few examples of cross-border transmission occurred, likely due to more effective non-pharmacological interventions applied at countries’ borders and that the major sink and source transmission pathways of SARS-CoV-2 with the Rio Grande do Sul state occurred between Brazilian states from the Southeast and Central West part of the country.

## 5. Conclusion

Overall, our study underscores the importance of robust sampling strategies and comprehensive genomic surveillance efforts to accurately track the spatio-temporal spread of SARS-CoV-2 lineages. By applying a sampling strategy based on the number of genomes and epidemiological information, we described the most prevalent clades of Gamma, Delta, and Omicron lineages that circulated in Rio Grande do Sul Brazil, during the period from June 2020 to July 2022 and showed that the large majority of transmission events took place between Brazilian states of the South-East and Central-West regions than between countries sharing land borders with Rio Grande do Sul.

## Data Availability

Data Availability The in-house script used to generate the study plots as well as the sampling numbers of each VOC in each brazilian state is publicly available here: https://github.com/dezordi/sc2_rs_2024. The gist tool developed in this study to create subsampling alignments using Augur and BLAST is publicly available here: https://github.com/dezordi/gist. The code lines used to conduct this study is publicly available in an online notebook: https://benchling.com/s/etr-VBPxhOEYsn5XG0g9W1tF?m=slm-u7Y y8Fp767dSSfHwFKrx.

## Acknowledgments

AGB and TFR thank FAPERGS (grant ARD #21/2551-0000703-6) for financial support. G.L.W is supported by the Conselho Nacional de Desenvolvimento Científico e Tecnológico (CNPq) through their productivity grant (307209/2023-7).

## References

1. A new coronavirus associated with human respiratory disease in China | Nature [Internet]. [cited 2024 Apr 22]. Available from: https://www.nature.com/articles/s41586-020-2008-3

2. WHO Director-General’s opening remarks at the media briefing on COVID-19 - 11 March 2020 [Internet]. [cited 2024 Apr 22]. Available from: https://www.who.int/director-general/speeches/detail/who-director-general-s-opening-remarks-at-the-media-briefing-on-covid-1911-march-2020

3. Johns Hopkins Coronavirus Resource Center [Internet]. [cited 2024 Apr 22]. COVID-19 Map. Available from: https://coronavirus.jhu.edu/map.html

4. Yanes-Lane M, Winters N, Fregonese F, Bastos M, Perlman-Arrow S, Campbell JR, et al. Proportion of asymptomatic infection among COVID-19 positive persons and their transmission potential: A systematic review and meta-analysis. PLOS ONE. 2020 Nov 3;15(11):e0241536.

5. Hu Z, Song C, Xu C, Jin G, Chen Y, Xu X, et al. Clinical characteristics of 24 asymptomatic infections with COVID-19 screened among close contacts in Nanjing, China. Sci China Life Sci. 2020 May 1;63(5):706–11.

6. Harrison AG, Lin T, Wang P. Mechanisms of SARS-CoV-2 Transmission and Pathogenesis. Trends Immunol. 2020 Dec 1;41(12):1100–15.

7. CDC. Centers for Disease Control and Prevention. 2020 [cited 2024 Apr 22]. Coronavirus Disease 2019 (COVID-19). Available from: https://www.cdc.gov/coronavirus/2019-ncov/variants/variant-classifications.html

8. Vaccines – COVID19 Vaccine Tracker [Internet]. [cited 2024 Apr 22]. Available from: https://covid19.trackvaccines.org/vaccines/#approved

9. Mohammed I, Nauman A, Paul P, Ganesan S, Chen KH, Jalil SMS, et al. The efficacy and effectiveness of the COVID-19 vaccines in reducing infection, severity, hospitalization, and mortality: a systematic review. Hum Vaccines Immunother [Internet]. 2022 Jan 31 [cited 2024 Apr 22]; Available from: https://www.tandfonline.com/doi/abs/10.1080/21645515.2022.2027160

10. Thompson Mark G., Stenehjem Edward, Grannis Shaun, Ball Sarah W., Naleway Allison L., Ong Toan C., et al. Effectiveness of Covid-19 Vaccines in Ambulatory and Inpatient Care Settings. N Engl J Med. 2021 Oct 6;385(15):1355–71.

11. Anand P, Stahel VP. The safety of Covid-19 mRNA vaccines: a review. Patient Saf Surg. 2021 May 1;15(1):20.

12. Dejnirattisai W, Zhou D, Supasa P, Liu C, Mentzer AJ, Ginn HM, et al. Antibody evasion by the P.1 strain of SARS-CoV-2. Cell. 2021 May 27;184(11):2939-2954.e9.

13. Hoffmann M, Arora P, Groß R, Seidel A, Hörnich BF, Hahn AS, et al. SARS-CoV-2 variants B.1.351 and P.1 escape from neutralizing antibodies. Cell. 2021 Apr 29;184(9):2384-2393.e12.

14. Ai J, Zhang H, Zhang Y, Lin K, Zhang Y, Wu J, et al. Omicron variant showed lower neutralizing sensitivity than other SARS-CoV-2 variants to immune sera elicited by vaccines after boost. Emerg Microbes Infect. 2022 Dec 31;11(1):337–43.

15. Hill V, Du Plessis L, Peacock TP, Aggarwal D, Colquhoun R, Carabelli AM, et al. The origins and molecular evolution of SARS-CoV-2 lineage B.1.1.7 in the UK. Virus Evol. 2022 Jul 1;8(2):veac080.

16. Tegally H, Wilkinson E, Giovanetti M, Iranzadeh A, Fonseca V, Giandhari J, et al. Detection of a SARS-CoV-2 variant of concern in South Africa. Nature. 2021 Apr;592(7854):438–43.

17. Naveca FG, Nascimento V, de Souza VC, Corado A de L, Nascimento F, Silva G, et al. COVID-19 in Amazonas, Brazil, was driven by the persistence of endemic lineages and P.1 emergence. Nat Med. 2021 Jul;27(7):1230–8.

18. McCrone JT, Hill V, Bajaj S, Pena RE, Lambert BC, Inward R, et al. Context-specific emergence and growth of the SARS-CoV-2 Delta variant. Nature. 2022 Oct;610(7930):154–60.

19. Viana R, Moyo S, Amoako DG, Tegally H, Scheepers C, Althaus CL, et al. Rapid epidemic expansion of the SARS-CoV-2 Omicron variant in southern Africa. Nature. 2022 Mar;603(7902):679–86.

20. Tracking SARS-CoV-2 variants [Internet]. [cited 2024 Apr 22]. Available from: https://www.who.int/activities/tracking-SARS-CoV-2-variants

21. Brito AF, Semenova E, Dudas G, Hassler GW, Kalinich CC, Kraemer MUG, et al. Global disparities in SARS-CoV-2 genomic surveillance. Nat Commun. 2022 Nov 16;13(1):7003.

22. Robishaw JD, Alter SM, Solano JJ, Shih RD, DeMets DL, Maki DG, et al. Genomic surveillance to combat COVID-19: challenges and opportunities. Lancet Microbe. 2021 Sep 1;2(9):e481–4.

23. Faria NR, Mellan TA, Whittaker C, Claro IM, Candido D da S, Mishra S, et al. Genomics and epidemiology of the P.1 SARS-CoV-2 lineage in Manaus, Brazil. Science. 2021 May 21;372(6544):815–21.

24. Giovanetti M, Fonseca V, Wilkinson E, Tegally H, San EJ, Althaus CL, et al. Replacement of the Gamma by the Delta variant in Brazil: Impact of lineage displacement on the ongoing pandemic. Virus Evol. 2022 Jan 1;8(1):veac024.

25. Giovanetti M, Slavov SN, Fonseca V, Wilkinson E, Tegally H, Patané JSL, et al. Genomic epidemiology of the SARS-CoV-2 epidemic in Brazil. Nat Microbiol. 2022 Sep;7(9):1490–500.

26. Nonaka CKV, Franco MM, Gräf T, Barcia CA de L, Mendonça RN de Á, Sousa KAF de, et al. Genomic Evidence of SARS-CoV-2 Reinfection Involving E484K Spike Mutation, Brazil - Volume 27, Number 5—May 2021 - Emerging Infectious Diseases journal - CDC. [cited 2024 Apr 22]; Available from: https://wwwnc.cdc.gov/eid/article/27/5/21-0191_article

27. Arantes I, Gomes Naveca F, Gräf T, COVID-19 Fiocruz Genomic Surveillance Network, Miyajima F, Faoro H, et al. Emergence and Spread of the SARS-CoV-2 Variant of Concern Delta across Different Brazilian Regions. Microbiol Spectr. 2022 Aug 24;10(5):e02641–21.

28. Mayer A de M, Gröhs Ferrareze PA, de Oliveira LFV, Gregianini TS, Neves CLAM, Caldana GD, et al. Genomic characterization and molecular evolution of SARS-CoV-2 in Rio Grande do Sul State, Brazil. Virology. 2023 May;582:1–11.

29. Martins AF, Zavascki AP, Wink PL, Volpato FCZ, Monteiro FL, Rosset C, et al. Detection of SARS-CoV-2 lineage P.1 in patients from a region with exponentially increasing hospitalisation rate, February 2021, Rio Grande do Sul, Southern Brazil. Eurosurveillance. 2021 Mar 25;26(12):2100276.

30. Francisco Jr R da S, Benites LF, Lamarca AP, de Almeida LGP, Hansen AW, Gularte JS, et al. Pervasive transmission of E484K and emergence of VUI-NP13L with evidence of SARS-CoV-2 co-infection events by two different lineages in Rio Grande do Sul, Brazil. Virus Res. 2021 Apr 15;296:198345.

31. Franceschi VB, Caldana GD, Perin C, Horn A, Peter C, Cybis GB, et al. Predominance of the SARS-CoV-2 Lineage P.1 and Its Sublineage P.1.2 in Patients from the Metropolitan Region of Porto Alegre, Southern Brazil in March 2021. Pathogens. 2021 Aug;10(8):988.

32. Franceschi VB, Caldana GD, de Menezes Mayer A, Cybis GB, Neves CAM, Ferrareze PAG, et al. Genomic epidemiology of SARS-CoV-2 in Esteio, Rio Grande do Sul, Brazil. BMC Genomics. 2021 May 20;22(1):371.

33. Quick J. nCoV-2019 sequencing protocol. 2020 Jan 22 [cited 2024 Apr 22]; Available from: https://www.protocols.io/view/ncov-2019-sequencing-protocol-bbmuik6w

34. Itokawa K, Sekizuka T, Hashino M, Tanaka R, Kuroda M. Disentangling primer interactions improves SARS-CoV-2 genome sequencing by multiplex tiling PCR. PLOS ONE. 2020 Sep 18;15(9):e0239403.

35. Naveca FG, Nascimento V, Souza V, Corado A de L, Nascimento F, Silva G, et al. Spread of Gamma (P.1) Sub-Lineages Carrying Spike Mutations Close to the Furin Cleavage Site and Deletions in the N-Terminal Domain Drives Ongoing Transmission of SARS-CoV-2 in Amazonas, Brazil. Microbiol Spectr. 2022 Feb 23;10(1):e02366–21.

36. Dezordi FZ, Neto AM da S, Campos T de L, Jeronimo PMC, Aksenen CF, Almeida SP, et al. ViralFlow: A Versatile Automated Workflow for SARS-CoV-2 Genome Assembly, Lineage Assignment, Mutations and Intrahost Variant Detection. Viruses. 2022 Feb;14(2):217.

37. Chen S, Zhou Y, Chen Y, Gu J. fastp: an ultra-fast all-in-one FASTQ preprocessor. Bioinformatics. 2018 Sep 1;34(17):i884–90.

38. Li H, Durbin R. Fast and accurate short read alignment with Burrows–Wheeler transform. Bioinformatics. 2009 Jul 15;25(14):1754–60.

39. Grubaugh ND, Gangavarapu K, Quick J, Matteson NL, De Jesus JG, Main BJ, et al. An amplicon-based sequencing framework for accurately measuring intrahost virus diversity using PrimalSeq and iVar. Genome Biol. 2019 Jan 8;20(1):8.

40. shiquan. shiquan/bamdst [Internet]. 2024 [cited 2024 Apr 22]. Available from: https://github.com/shiquan/bamdst

41. O’Toole Á, Scher E, Underwood A, Jackson B, Hill V, McCrone JT, et al. Assignment of epidemiological lineages in an emerging pandemic using the pangolin tool. Virus Evol. 2021;7(2):veab064.

42. De Maio N, Kalaghatgi P, Turakhia Y, Corbett-Detig R, Minh BQ, Goldman N. Maximum likelihood pandemic-scale phylogenetics. Nat Genet. 2023 May;55(5):746–52.

43. Khare S, Gurry C, Freitas L, Schultz MB, Bach G, Diallo A, et al. GISAID’s Role in Pandemic Response. China CDC Wkly. 2021 Dec 3;3(49):1049–51.

44. Huddleston J, Hadfield J, Sibley TR, Lee J, Fay K, Ilcisin M, et al. Augur: a bioinformatics toolkit for phylogenetic analyses of human pathogens. J Open Source Softw. 2021 Jan 7;6(57):2906.

45. Altschul SF, Gish W, Miller W, Myers EW, Lipman DJ. Basic local alignment search tool. J Mol Biol. 1990 Oct 5;215(3):403–10.

46. Katoh K, Standley DM. MAFFT Multiple Sequence Alignment Software Version 7: Improvements in Performance and Usability. Mol Biol Evol. 2013 Apr 1;30(4):772–80.

47. Minh BQ, Schmidt HA, Chernomor O, Schrempf D, Woodhams MD, von Haeseler A, et al. IQ-TREE 2: New Models and Efficient Methods for Phylogenetic Inference in the Genomic Era. Mol Biol Evol. 2020 May 1;37(5):1530–4.

48. Kalyaanamoorthy S, Minh BQ, Wong TKF, von Haeseler A, Jermiin LS. ModelFinder: fast model selection for accurate phylogenetic estimates. Nat Methods. 2017 Jun;14(6):587–9.

49. Guindon S, Dufayard JF, Lefort V, Anisimova M, Hordijk W, Gascuel O. New Algorithms and Methods to Estimate Maximum-Likelihood Phylogenies: Assessing the Performance of PhyML 3.0. Syst Biol. 2010 May 1;59(3):307–21.

50. Letunic I, Bork P. Interactive Tree Of Life (iTOL): an online tool for phylogenetic tree display and annotation. Bioinforma Oxf Engl. 2007 Jan 1;23(1):127–8.

51. Rambaut A, Lam TT, Max Carvalho L, Pybus OG. Exploring the temporal structure of heterochronous sequences using TempEst (formerly Path-O-Gen). Virus Evol. 2016 Jan 1;2(1):vew007.

52. Sagulenko P, Puller V, Neher RA. TreeTime: Maximum-likelihood phylodynamic analysis. Virus Evol. 2018 Jan 1;4(1):vex042.

53. Morel B, Barbera P, Czech L, Bettisworth B, Hübner L, Lutteropp S, et al. Phylogenetic Analysis of SARS-CoV-2 Data Is Difficult. Mol Biol Evol. 2021 May 4;38(5):1777–91.

54. Hill V, Ruis C, Bajaj S, Pybus OG, Kraemer MUG. Progress and challenges in virus genomic epidemiology. Trends Parasitol. 2021 Dec 1;37(12):1038–49.

55. Mir D, Rego N, Resende PC, Tort F, Fernández-Calero T, Noya V, et al. Recurrent Dissemination of SARS-CoV-2 Through the Uruguayan–Brazilian Border. Front Microbiol [Internet]. 2021 May 28 [cited 2024 Apr 22];12. Available from: https://www.frontiersin.org/journals/microbiology/articles/10.3389/fmicb.2021.653986/full

56. Ferrareze PAG, Cybis GB, de Oliveira LFV, Zimerman RA, Schiavon DEB, Peter C, et al. Intense P.1 (Gamma) diversification followed by rapid Delta substitution in Southern Brazil: a SARS-CoV-2 genomic epidemiology study. Microbes Infect. 2024 Jan 1;26(1):105216.

57. Y Castro TR, Piccoli BC, Vieira AA, Casarin BC, Tessele LF, Salvato RS, et al. Introduction, Dispersal, and Predominance of SARS-CoV-2 Delta Variant in Rio Grande do Sul, Brazil: A Retrospective Analysis. Microorganisms. 2023 Dec 7;11(12):2938.

58. da Silva MS, Gularte JS, Filippi M, Demoliner M, Girardi V, Mosena ACS, et al. Genomic and epidemiologic surveillance of SARS-CoV-2 in Southern Brazil and identification of a new Omicron-L452R sublineage. Virus Res. 2022 Nov;321:198907.

59. Rhoden J, Hoffmann AT, Stein JF, da Silva MS, Gularte JS, Filippi M, et al. Diversity of Omicron sublineages and clinical characteristics in hospitalized patients in the southernmost state of Brazil. BMC Infect Dis. 2024 Feb 13;24(1):193.

